# Dysplasia-Stratified Management of Barrett’s Esophagus: An Incidence-Based U.S. Cost-Effectiveness Analysis

**DOI:** 10.64898/2026.06.12.26355512

**Authors:** Akiko Kowada

## Abstract

**Background and Aims:** Barrett’s esophagus (BE) is the principal precursor of esophageal adenocarcinoma (EAC), whose incidence has risen sharply in Western countries since the 1960s. Effective, dysplasia-stratified surveillance strategies are needed to prevent progression. This study evaluated the cost-effectiveness of dysplasia-stratified surveillance intervals and endoscopic eradication therapy (EET) across the BE spectrum.

**Methods:** We developed an incidence-based Markov state-transition model of BE progression calibrated to U.S. epidemiologic data from a healthcare-sector perspective over a lifetime horizon. Four hypothetical cohorts of 50-year-old individuals with short-segment BE (SSBE), nondysplastic BE (NDBE), low-grade dysplasia (LGD), or high-grade dysplasia (HGD) were evaluated. Strategies included no surveillance; surveillance at 1-, 2-, 3-, 4-, 5-, or 10-year intervals; standard or AI-assisted endoscopy; non-endoscopic screening (sponge, breath, miRNA tests); and EET for LGD and HGD. Outcomes included costs, quality-adjusted life years (QALYs), incremental cost-effectiveness ratios (ICERs), net monetary benefits (NMBs), EAC cases, and EAC-related deaths. Sensitivity analyses used a willingness-to-pay threshold of US$100,000 per QALY.

**Results:** No surveillance was the most cost-effective strategy for SSBE and NDBE. For LGD, upfront EET was more cost-effective than all surveillance strategies, with results sensitive to EAC incidence and recurrence. For HGD, EET was cost-saving and yielded the greatest QALYs, with findings robust in 99.9% of simulations. EET prevented 12,614 and 44,295 EAC-related deaths per 100,000 individuals with LGD and HGD, respectively.

**Conclusion:** Dysplasia-stratified management is essential for optimizing surveillance and treatment strategies in BE. Any degree of dysplasia should receive EET followed by targeted post-treatment monitoring, establishing EET as the central therapeutic pathway for dysplastic BE.

## 1. Introduction

Barrett’s esophagus (BE) is the principal precursor to esophageal adenocarcinoma (EAC) [1], a malignancy whose incidence has risen more rapidly than any other esophageal epithelial cancer [2,3]. Globally, EAC incidence has increased steadily over recent decades, with the highest rates observed in North America and Western Europe. In the United States, incidence has risen more than seven-fold, and the 5-year survival for advanced-stage disease remains below 20%. Paradoxically, recent epidemiologic analyses show a decline in early-stage EAC detection, suggesting that current surveillance strategies may be insufficient to intercept progression at a curable stage [4].

Dysplasia grade is the strongest predictor of progression from BE to EAC. In a large U.S. cohort, 70.1% of individuals with BE had nondysplastic BE (NDBE), 11.5% had low-grade dysplasia (LGD), and 5.4% had high-grade dysplasia (HGD) [5]. Updated incidence estimates from a recent meta-analysis demonstrate annual progression rates of 0.21% for NDBE, 1.16% for LGD, and 14.16% for HGD, underscoring the markedly heterogeneous natural history of BE [6]. Progression risk is further influenced by segment length and patient-level factors such as sex, obesity, smoking, and reflux severity [7,8]. These data support a dysplasia-stratified approach to surveillance and management.

Despite this heterogeneity, current U.S. guidelines recommend uniform surveillance intervals for NDBE (every 3–5 years) and endorse endoscopic eradication therapy (EET) for LGD and HGD [9]. These recommendations are largely based on European evidence and do not incorporate recent U.S. data questioning the effectiveness and cost-effectiveness of NDBE surveillance, including randomized trials showing no reduction in EAC-related mortality and economic analyses suggesting poor value in low-risk populations [10]. As a result, the optimal management of LGD and the value of continued surveillance for NDBE in the U.S. healthcare system remain uncertain.

Meanwhile, advances in AI-assisted endoscopy [11,12] and non-endoscopic screening technologies, including sponge-based, breath-based, and molecular assays, are reshaping the landscape of BE detection and surveillance [13,14,15]. These developments highlight the need to re-evaluate existing surveillance paradigms and determine whether dysplasia-stratified strategies can more efficiently prevent EAC while minimizing unnecessary procedures and optimizing resource allocation.

To inform evidence-based surveillance policy in the United States, the cost-effectiveness of dysplasia-stratified surveillance and EET strategies requires urgent evaluation. The aim of this study is to assess the cost-effectiveness of dysplasia-stratified EAC surveillance and treatment strategies across varying surveillance intervals in the U.S. healthcare system.

## 2. Methods

### 2.1 Model Overview

We developed an incidence-based Markov state-transition model simulating the natural history of BE across four phenotypes: short-segment BE (SSBE), NDBE, LGD, and HGD (Figure 1). Each phenotype was modeled as an independent cohort to reflect phenotype-specific annual incidence rates of EAC.

**Figure 1.**
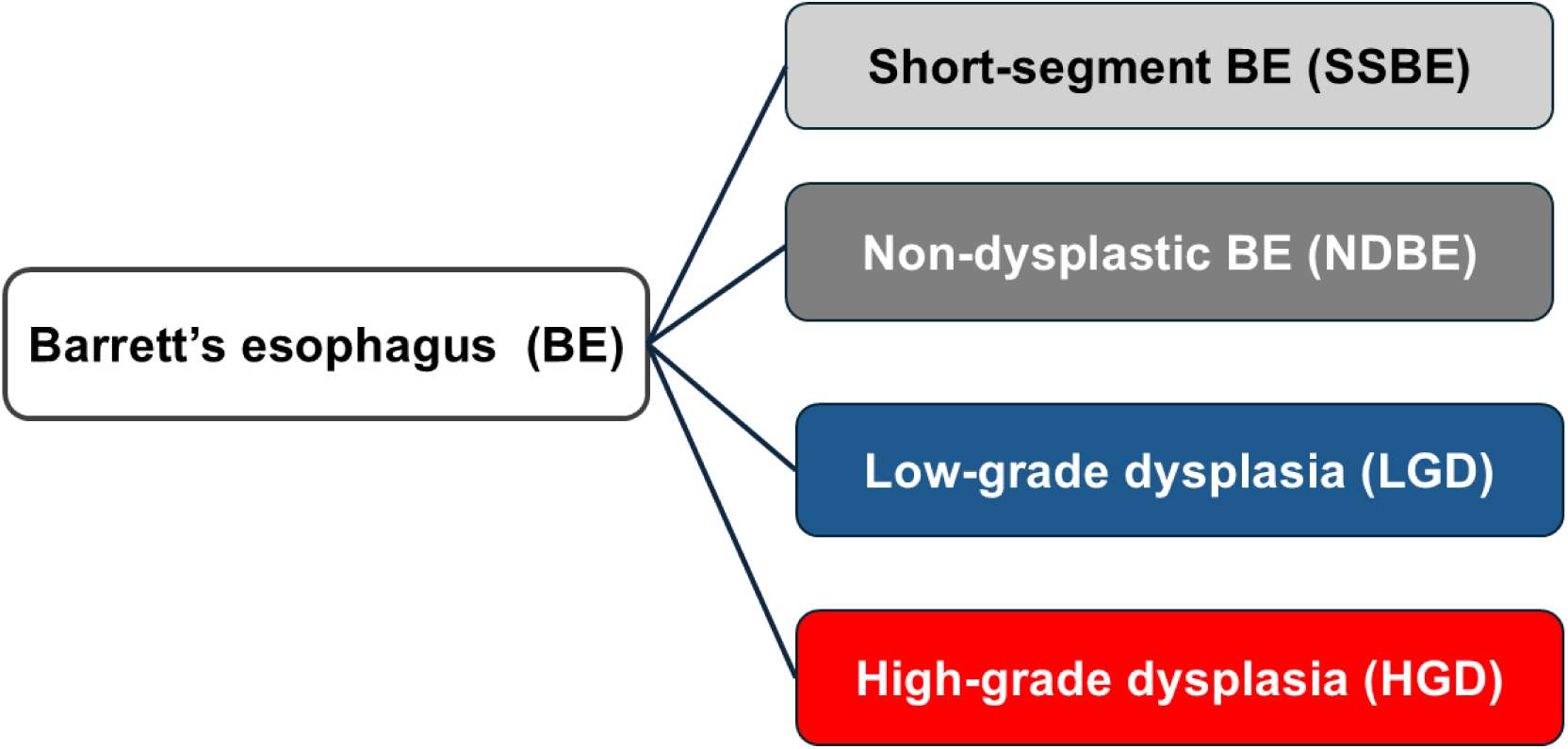
Classification of Barrett’s esophagus phenotypes.

The model was calibrated to contemporary U.S. epidemiologic data and conducted from a healthcare-sector perspective over a lifetime horizon, using annual cycles with 3% discounting for both costs and quality-adjusted life-years (QALYs) [16]. A half-cycle correction was applied. Age 50 was selected because it approximates the median age at BE diagnosis and corresponds to the period when EAC risk begins to rise.

Five surveillance strategies were evaluated for each phenotype:

1. No surveillance
2. Standard endoscopic surveillance
3. AI-assisted endoscopic surveillance
4. Non-endoscopic surveillance (sponge, breath, miRNA tests)
5. Upfront EET (LGD and HGD only)

Surveillance intervals of 1, 2, 3, 4, 5, and 10 years were assessed. Positive non-endoscopic tests triggered confirmatory endoscopy, whereas individuals with negative results continued surveillance at the assigned interval. Individuals undergoing upfront EET received therapeutic endoscopy followed by scheduled post-treatment endoscopic assessment.

### 2.2 Model Structure

Four independent Markov models were constructed to reflect phenotype-specific EAC incidence. Individuals remained in their assigned BE state unless they progressed to EAC, died, or underwent EET, after which they transitioned to the corresponding cancer or post-treatment state (Figure 2). Transitions between BE phenotypes were not modeled because the analysis relied on observed EAC incidence and mortality rather than hypothetical natural-history progression pathways.

**Figure 2.**
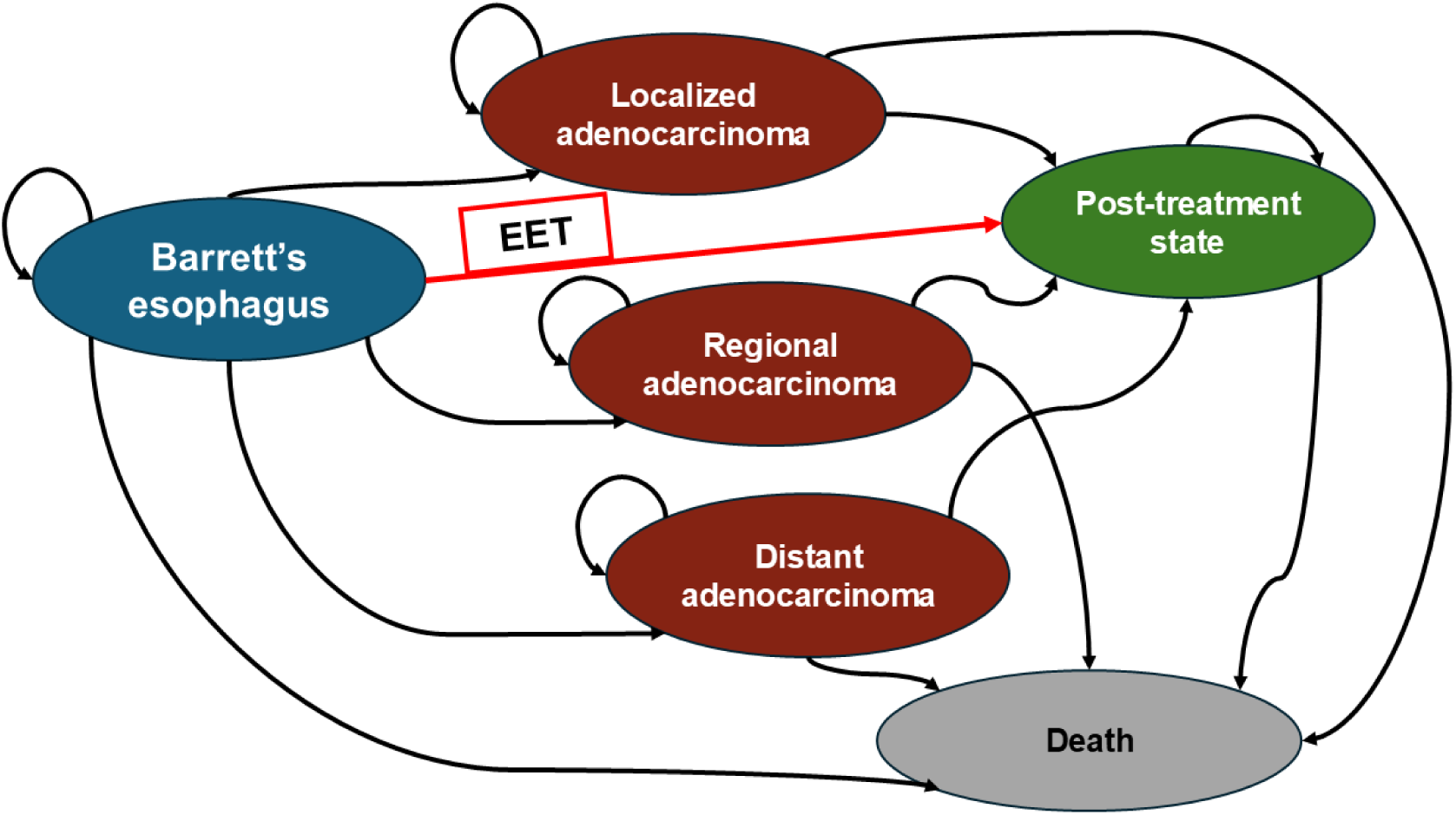
State-transition diagram for Barrett’s esophagus progression. The model does not simulate transitions between cancer stages; instead, each cancer stage is modeled as an independent incident state. Arrows represent conceptual pathways of disease progression, treatment with EET, and mortality. EET, endoscopic eradication therapy.

All EAC cases were assumed to occur and be diagnosed within the same annual cycle according to phenotype-specific incidence rates. Recurrence of dysplasia and EAC after EET was incorporated for LGD and HGD. Mortality included stage-specific EAC survival and background mortality derived from SEER life tables.

Model outcomes included total costs, QALYs, incremental cost-effectiveness ratios (ICERs), net monetary benefits (NMBs), and the number of EAC cases and EAC-related deaths.

### 2.3 Model Inputs

Clinical probabilities and epidemiologic parameters were obtained through a MEDLINE search (2000–2026) and from Surveillance, Epidemiology, and End Results (SEER) cancer statistics (Supplementary Table S1). Annual EAC incidence ranged from 0.0006 for SSBE to 0.1416 for HGD [6,17]. Recurrence of dysplasia and EAC after EET was incorporated for LGD and HGD [18,19]. Adherence to surveillance ranged from 0.78 to 0.95 [20], with one-way sensitivity analyses varying adherence from 0.50 to 1.00. Sensitivity and specificity for each surveillance modality were derived from published studies [9,12,13,14,15]. Stage distribution and stage-specific survival were calibrated to SEER data.

Costs included standard endoscopy [21], AI-assisted endoscopy, non-endoscopic tests [14,22], stage-specific EAC treatment [23], and EET for LGD and HGD [24]. Stage-specific EAC treatment costs were calibrated to SEER Medicare–based estimates from Tramontano et al. [23]. Costs for EET were derived from Medicare facility payment rates for relevant CPT codes (43254, 43270, 43239, 88305) in the 2026 CMS Physician Fee Schedule [24]. Parameters without available empirical evidence were based on reasonable assumptions and expert opinion, as detailed in Supplementary Table S1.

### 2.4 Health State Utilities

Health state utilities were assigned to each BE and esophageal cancer state. Utilities were applied to time spent in each health state without modeling annual transitions, consistent with the incidence-based structure of the model. Utility values were obtained from published literature [25] and discounted at 3% annually [16].

### 2.5 Sensitivity Analyses

One-way sensitivity analyses varied key clinical and economic parameters across plausible ranges, including test sensitivity and specificity, adherence to surveillance, surveillance and treatment costs, health-state utilities, and phenotype-specific EAC incidence and recurrence rates (Supplementary Table S1). Deterministic results were summarized using tornado diagrams.

A probabilistic sensitivity analysis (PSA) was conducted using 10,000 Monte Carlo simulations. Parameter uncertainty was represented using beta distributions for test performance, phenotype-specific EAC incidence, and health-state utilities; Dirichlet distributions for stage distribution; and gamma distributions for costs. For each simulation, lifetime costs and QALYs were calculated, and cost-effectiveness acceptability curves were generated at a willingness-to-pay (WTP) threshold of US$100,000 per QALY gained.

### 2.6 Scenario Analyses in the NDBE Cohort

Smoking and obesity were evaluated as risk factors for EAC. Odds ratios for ever smoking (OR = 1.96; 95% CI: 1.64–2.34) [7] and obesity (OR = 2.51; 95% CI: 1.54–4.06) [8] were obtained from the literature. Scenario analyses applied these risk-specific incidence rates to the model, and corresponding costs, QALYs, and ICERs were calculated for each scenario.

### 2.7 Markov Cohort Analyses

Using the Markov cohort model, we estimated cumulative lifetime numbers of localized, regional, and distant EAC cases; EAC-related deaths averted; cost savings; and QALY gains of EET compared with no surveillance. For the LGD and HGD cohorts, outcomes per 100,000 patients were calculated by multiplying the difference in cumulative probabilities between EET and no surveillance by 100,000.

All analyses were conducted using TreeAge Pro 2026 (TreeAge Software, Williamstown, MA). This economic evaluation followed the CHEERS 2022 reporting guidelines [26].

### 2.8 Model Validation

We performed face, internal, and external validation to ensure that the model structure, parameterization, and outputs were clinically plausible and consistent with established epidemiologic patterns.

Face validation assessed model structure, assumptions, and parameter inputs for alignment with established clinical pathways and prior decision-analytic frameworks in BE and EAC. Health states, recurrence patterns after EET, and phenotype-specific incidence inputs were evaluated for consistency with current clinical understanding.

Internal validation examined whether simulated transitions and long-term outcomes behaved as expected across the four BE phenotypes. Annual EAC incidence, stage distribution at diagnosis, recurrence after EET, and survival patterns were verified to reproduce known epidemiologic gradients across SSBE, NDBE, LGD, and HGD. Model behavior under extreme parameter values and across sensitivity analyses was assessed for logical consistency and numerical stability.

External validation compared model-generated estimates with independent epidemiologic data. Stage distribution and stage-specific survival were obtained directly from SEER, and annual EAC incidence and recurrence after EET were derived from published cohort studies. The model reproduced the expected incidence gradient from SSBE to HGD, the predominance of localized disease at diagnosis, and stage-specific survival patterns, supporting its suitability for evaluating dysplasia-stratified surveillance strategies in BE.

## 3. Results

### 3.1 Base-case results

In the base-case analysis, no surveillance was the most cost-effective strategy for SSBE and NDBE (Figure 3, Supplementary Table S2 and S3). Across these phenotypes, neither non-endoscopic surveillance nor endoscopic surveillance at any interval met the WTP threshold of US$100,000 per QALY gained.

**Figure 3.**
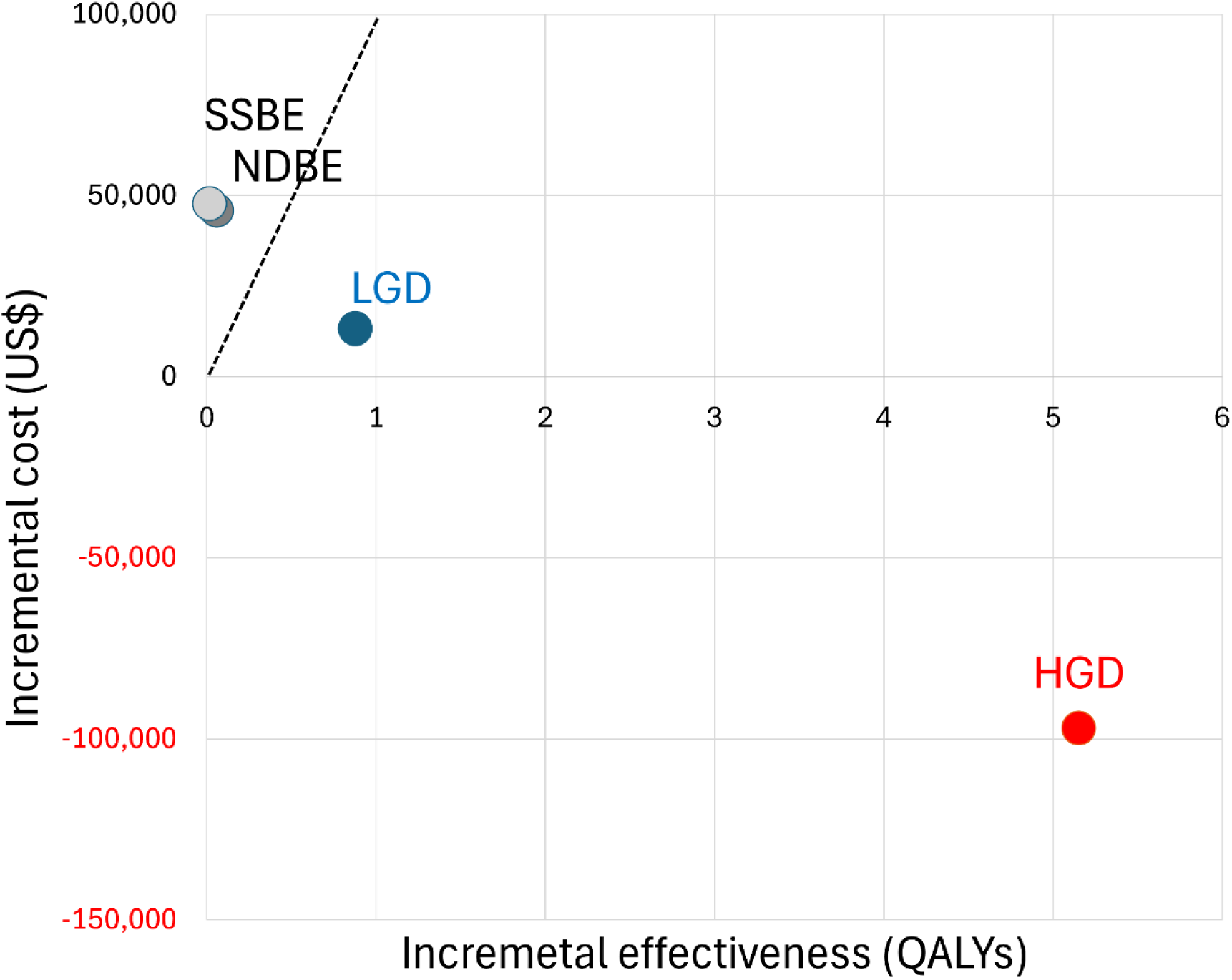
Cost-effectiveness plane in the LGD cohort. The dashed line represents the willingness-to-pay (WTP) threshold of $100,000 per QALY gained. SSBE, short-segment Barrett’s esophagus; NDBE, non-dysplastic Barrett’s esophagus; LGD, low-grade dysplasia; HGD, high-grade dysplasia.

For LGD, upfront EET was cost-effective, yielding US$64,234 in lifetime costs, 16.3790 QALYs, and an NMB of US$1,573,669, compared with US$51,169, 15.5039 QALYs, and an NMB of US$1,499,217 for no surveillance (Supplementary Table S4). For HGD, EET was cost-saving, with lower lifetime costs (US$115,450), higher QALYs (12.8171), and the highest NMB (US$1,166,264) (Supplementary Table S5). Surveillance-based strategies were consistently less efficient than EET for both LGD and HGD and did not outperform it at any interval.

### 3.2 Clinical outcomes, costs, and QALYs

EET produced substantial clinical benefits in the LGD and HGD cohorts. Per 100,000 individuals, EET detected 746 and 11,932 additional localized EAC cases, prevented 15,951 and 56,444 regional EAC cases, and prevented 18,131 and 60,668 distant EAC cases, respectively (Table 1). EET also averted 12,614 EAC-related deaths in LGD and 44,295 in HGD over a lifetime.

**Table 1.** Lifetime health outcomes of endoscopic eradication therapy among 100,000 individuals compared with no surveillance.

| BE state | Cost difference (US\$) | QALY gains (QALYs) | Localized EAC cases fewer detected (n) | Regional EAC cases prevented (n) | Distant EAC cases prevented (n) | EAC-related deaths averted (n) |
| --- | --- | --- | --- | --- | --- | --- |
| LGD | 1,306,416,972 | 87,516 | 746 | 15,951 | 18,131 | 12,614 |
| HGD | -9,703,459,221 | 515,299 | 11,932 | 56,444 | 60,668 | 44,295 |

BE, Barrett's esophagus; EAC, esophageal adenocarcinoma; LGD, low-grade dysplasia; HGD, high-grade dysplasia.

These clinical gains were accompanied by US$3.9 billion in additional costs for LGD and US$9.7 billion in cost savings for HGD, along with 87,516 and 515,299 additional QALYs, respectively (Table 1).

### 3.3 Sensitivity analyses

One-way sensitivity analyses in the LGD cohort showed that the cost-effectiveness of EET was driven primarily by phenotype-specific EAC incidence and recurrence. EET became cost-effective relative to no surveillance when the annual EAC incidence exceeded 0.007 and when recurrence to EAC was below 0.01 (Figure 4A).

**Figure 4.**
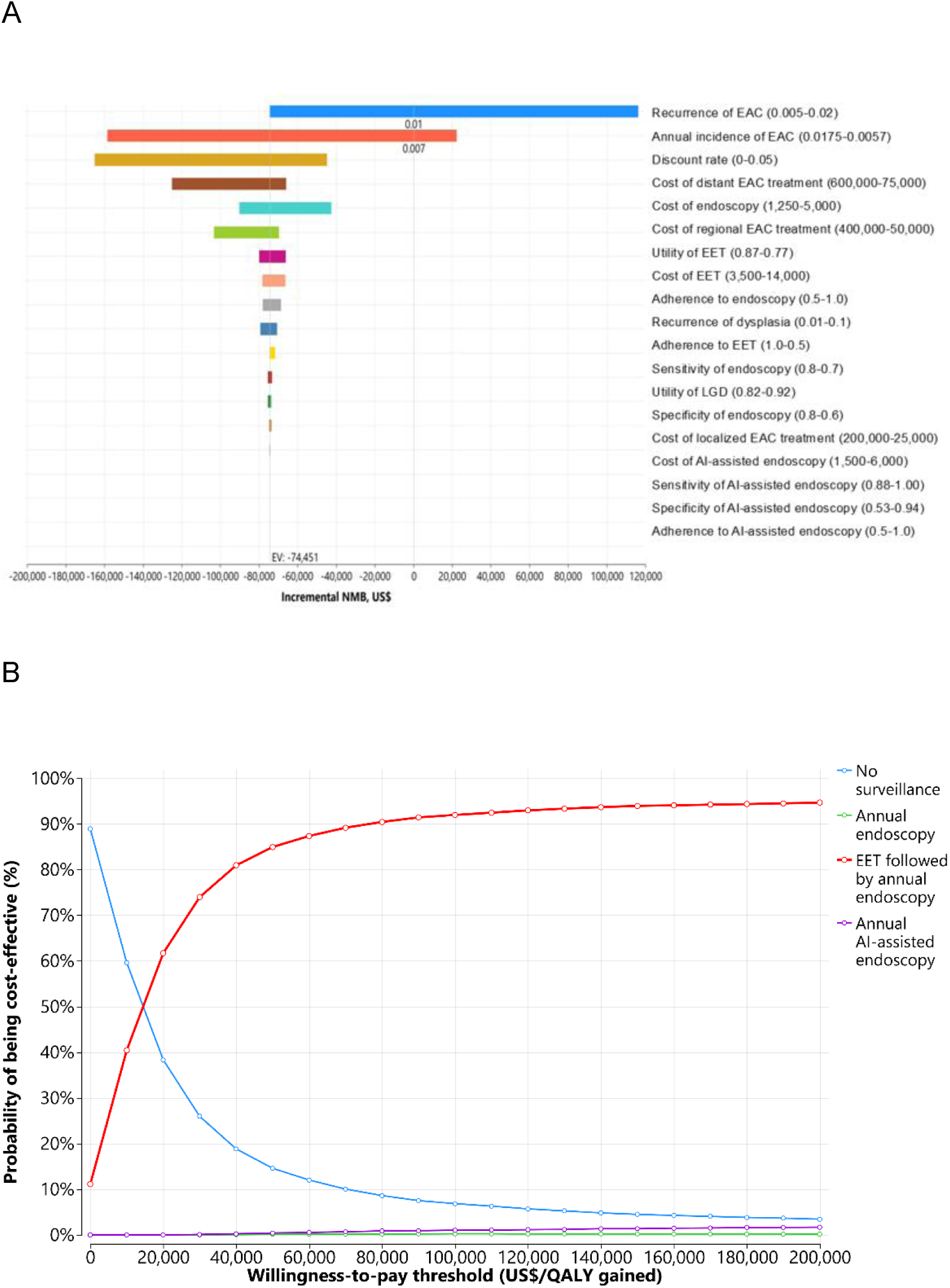
Sensitivity analyses in the LGD cohorts. (A) Tornado diagram comparing no surveillance with EET followed by annual endoscopy. (B) Cost-effectiveness acceptability curves. AI, artificial intelligence; EAC, esophageal adenocarcinoma; EET, endoscopic eradication therapy; LGD, low-grade dysplasia.

Cost-effectiveness acceptability curves demonstrated a 100% probability that no surveillance was cost-effective for SSBE and NDBE. Upfront EET had a 92.0% probability of being cost-effective for LGD at a WTP threshold of US$100,000 per QALY gained and remained highly robust for HGD, with a 99.9% probability of cost-effectiveness across probabilistic simulations (Figure 4B).

### 3.4 Scenario analyses in the NDBE cohort

Scenario analyses applying risk-specific incidence rates for smoking and obesity showed that annual AI-assisted endoscopy became the preferred strategy in the NDBE cohort when both risk factors were elevated. A two-way risk-surface analysis demonstrated a clear transition boundary, with annual AI-assisted endoscopy becoming optimal at approximately a smoking relative risk above 2.0 and an obesity relative risk above 3.0 (Figure 5).

**Figure 5.**
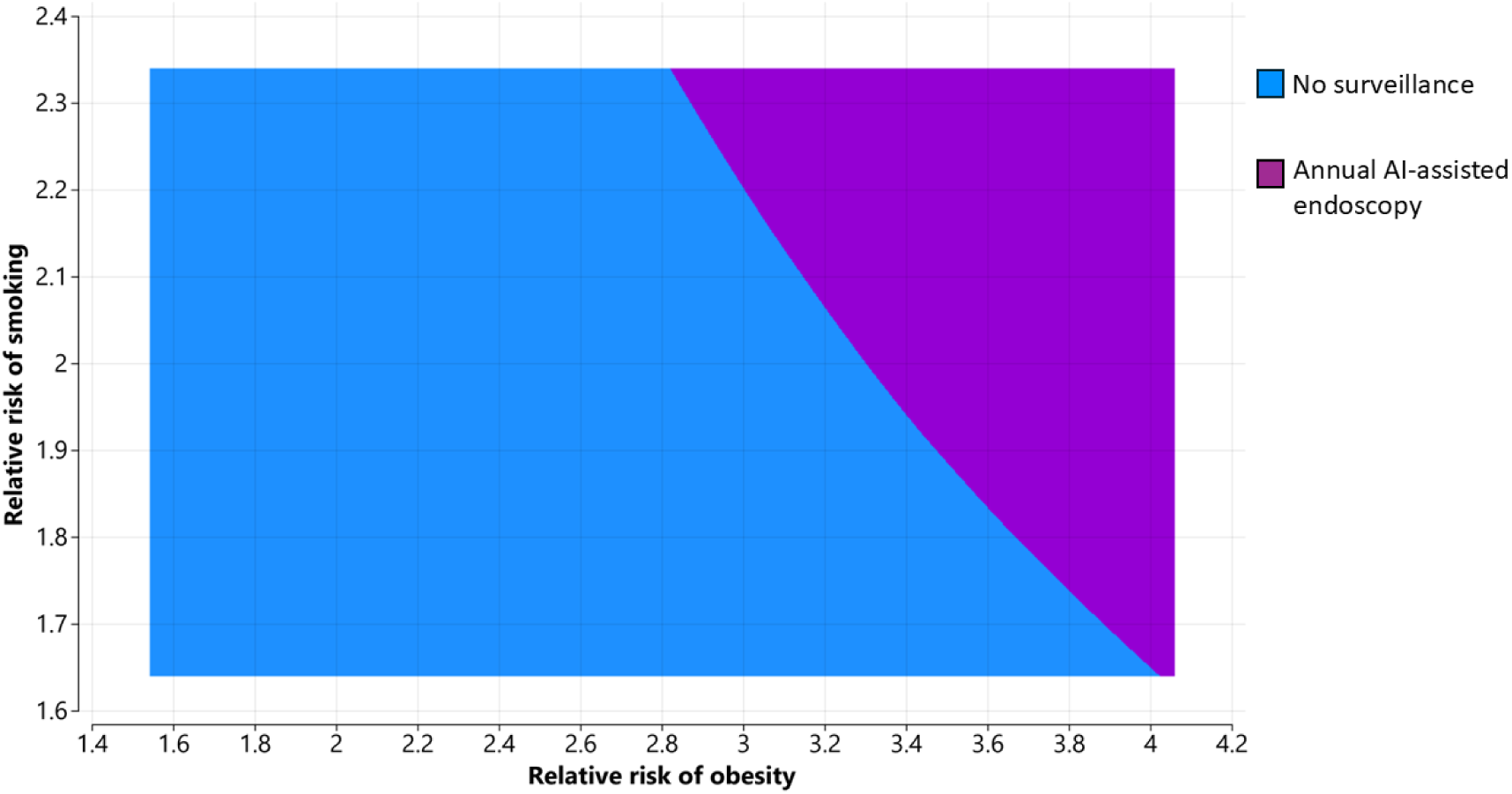
Two-way scenario analysis of smoking and obesity risks in the NDBE cohort. Colors indicate the optimal surveillance strategy for each risk combination: blue = no surveillance; purple = annual AI-assisted endoscopy

## 4. Discussion

### 4.1 Key Findings

In this comprehensive cost-effectiveness analysis across the full spectrum of BE, we found that dysplasia-stratified management yields markedly different value depending on phenotype. No surveillance was the most cost-effective strategy for individuals with SSBE or NDBE, whereas upfront EET was cost-effective for LGD and cost-saving for HGD. These findings demonstrate that applying uniform surveillance intervals across heterogeneous BE phenotypes is inefficient and may misallocate healthcare resources, while offering limited benefit for EAC prevention.

### 4.2 Interpretation

The divergent results across BE phenotypes reflect the steep gradient in EAC incidence. Recent meta-analytic estimates confirm that SSBE and NDBE carry extremely low annual risks of progression [6,17], making surveillance unlikely to generate meaningful QALY gains. In contrast, LGD and HGD exhibit substantially higher progression risks, and EET directly removes dysplastic epithelium, thereby interrupting the dysplasia-to-cancer sequence [18,19,27,28]. This mechanism explains why EET not only improved clinical outcomes but also produced large QALY gains and substantial cost savings in HGD.

Our study showed that the magnitude of clinical benefit was striking: HGD, EET prevented more than three times as many EAC-related deaths and detected sixteen times more localized cancers compared with LGD. These mortality reductions are driven by large shifts from regional and distant EAC to localized disease, consistent with the established effectiveness of EET in halting progression before invasion occurs.

### 4.3 Comparison With Prior Studies

Several cost-effectiveness analyses have previously evaluated surveillance strategies for BE. Kastelein et al. reported that surveillance every 5 years for NDBE and every 3 years for LGD was cost-effective in the Netherlands, assuming endoscopic therapy for HGD or early EAC and esophagectomy for advanced cancer, using a willingness-to-pay threshold of €35,000 per QALY [29]. Vissapragada et al. similarly identified risk-stratified surveillance as optimal in Australia, with biennial surveillance for long-segment BE (>2 cm) and 12-month surveillance for LGD providing the greatest value [30]. Omidvari et al. conducted a comparative modeling analysis using three National Cancer Institute CISNET esophageal adenocarcinoma models (MISCAN-EAC, EACMo, and MSCE-EAC) to evaluate 78 management strategies for NDBE and LGD [31]. They concluded that the optimal management strategy for patients with BE and LGD is EET, but only after LGD is confirmed by repeat endoscopy following high-dose acid suppression. For patients with NDBE, the most cost-effective strategy is endoscopic surveillance, using a 3-year interval for men and a 5-year interval for women. For patients with NDBE, the most cost-effective strategy is endoscopic surveillance, using a 3-year interval for men and a 5-year interval for women. Federici et al. evaluated LGD and HGD in an Italian Markov model and found that EET was cost-effective for LGD and dominant for HGD [32], consistent with our findings, although their analysis was limited to dysplasia and relied on older natural-history parameters.

The differences between their conclusions and ours arise not from small variations in individual parameters, but from fundamentally different modeling frameworks. All prior analyses relied on natural-history transition probabilities that are unobservable and require strong assumptions. In contrast, our model uses contemporary phenotype-specific incidence estimates [6], avoiding reliance on uncertain natural-history inputs. Whereas previous studies evaluated only portions of the BE spectrum, typically NDBE and LGD, we conducted cost-effective analyses for the full BE spectrum within a unified framework.

To our knowledge, our study is the first to evaluate the full BE spectrum using contemporary phenotype-specific incidence estimates. By incorporating natural-history-free, incidence-based inputs across all phenotypes, our model reflects the modern epidemiology of BE and enables direct comparison of surveillance and treatment strategies across SSBE, NDBE, LGD, and HGD. This approach clarifies, for the first time, the sharp divergence in value across phenotypes: no surveillance is cost-effective for SSBE and NDBE, whereas EET is consistently optimal for LGD and HGD. These findings also highlight the importance of sustained EAC prevention through effective initiation and maintenance of EET in dysplastic disease.

These findings support a simplified and actionable management paradigm for BE. Individuals with SSBE or NDBE may safely avoid routine surveillance, reducing unnecessary procedures and healthcare expenditures without compromising cancer prevention. In contrast, any degree of dysplasia should prompt EET, followed by limited post-treatment surveillance to monitor for recurrence. This dysplasia-directed approach has the potential to reduce EAC mortality while improving the efficiency of BE management in the United States. Our results also suggest that current ASGE guidelines may recommend surveillance more broadly than warranted in low-risk populations and underemphasize definitive therapy for dysplasia.

### 4.4 Scenario Analyses and Personalized Risk

In the NDBE cohort, smoking and obesity substantially increased EAC risk, and annual AI-assisted endoscopy became cost-effective in individuals with both factors elevated. These findings highlight the importance of incorporating individualized risk factors into surveillance decisions rather than relying solely on histologic phenotype. As AI-assisted endoscopy continues to improve in accuracy and accessibility, its role in personalized, risk-adapted surveillance may expand further, offering a scalable approach for identifying high-risk individuals who stand to benefit most from intensified monitoring [11,12].

### 4.5 Strengths and Limitations

Strengths of this study include the use of contemporary phenotype-specific incidence rates, incorporation of recurrence after EET, evaluation of multiple modern surveillance modalities, and extensive sensitivity and scenario analyses. The modeling framework used here has been applied previously to assess cancer-prevention strategies in several disease settings, including *Helicobacter pylori* eradication for gastric cancer and screening strategies for hepatocellular carcinoma, demonstrating its flexibility and validity across distinct prevention contexts [33,34,35]. The recent availability of phenotype-specific incidence estimates for BE now enables this framework to be applied to the full BE spectrum for the first time.

Limitations include uncertainty in dysplasia-specific incidence estimates, potential variation in real-world adherence, and assumptions inherent to state-transition modeling. Nonetheless, the model reproduced known epidemiologic gradients and survival patterns, supporting its validity for evaluating dysplasia-stratified surveillance strategies.

### 4.6 Future Directions

Future efforts to strengthen EAC prevention should build on the dysplasia-stratified framework highlighted in this analysis. For patients with low-and high-grade dysplasia, timely and appropriate use of EET represents the most effective strategy for reducing cancer risk, underscoring the need to optimize EET selection, delivery, and long-term post-treatment management. Although EET is highly effective in suppressing dysplasia progression, recurrent BE and dysplasia remain common, reflecting persistent biological susceptibility and ongoing reflux-related injury. These observations indicate that the natural history of BE continues even after apparently successful eradication, suggesting that long-term maintenance strategies may be required to achieve durable suppression of disease progression. In addition to endoscopic maintenance therapy, lifestyle-based risk-reduction measures such as weight loss, smoking cessation, and control of reflux-promoting behaviors may further reduce residual progression risk after EET. Figure 6 conceptually illustrates how sustained maintenance therapy could further suppress the natural history of BE beyond the effect of a single EET intervention.

**Figure 6.**
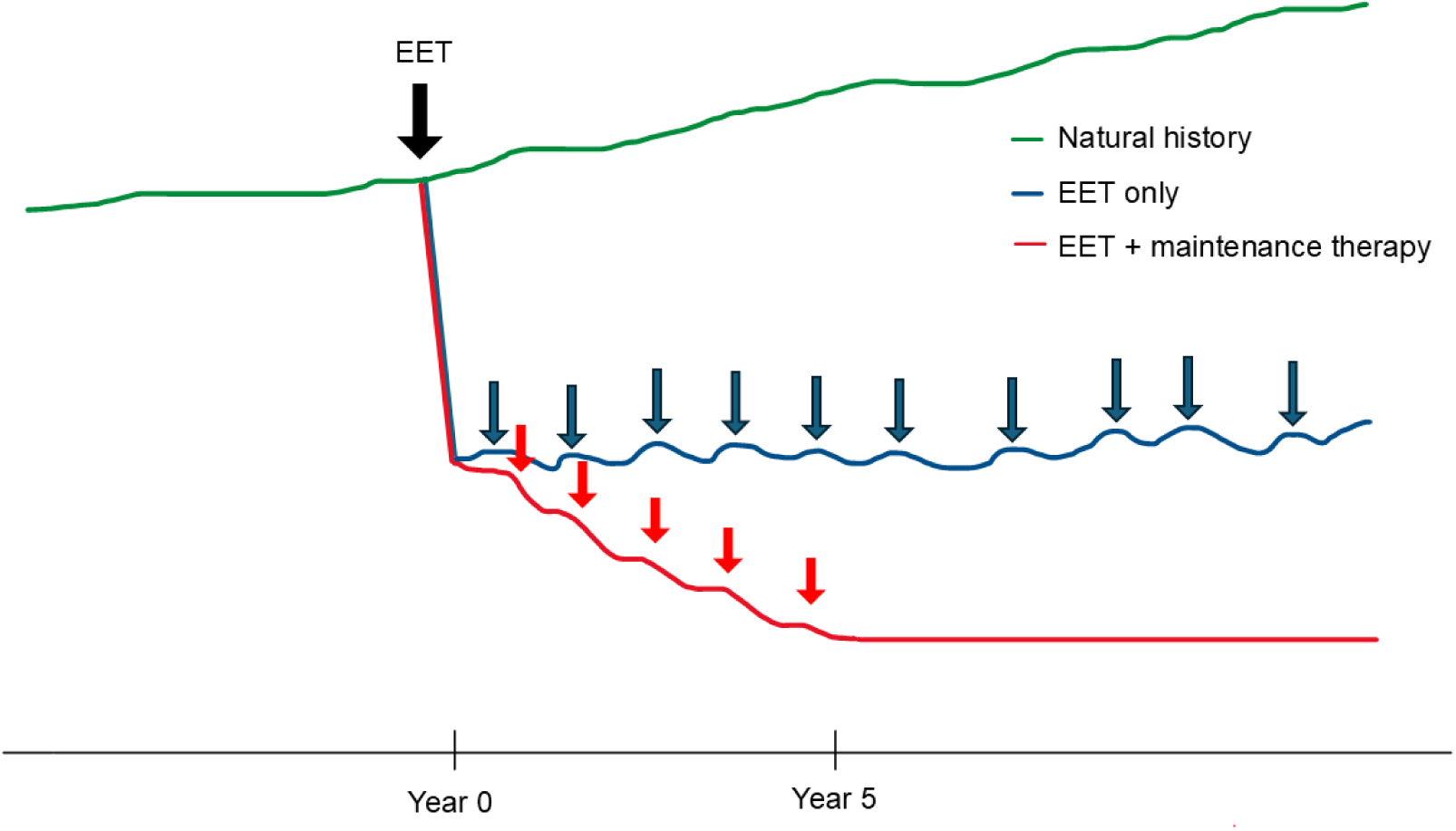
Conceptual model of EAC risk trajectories under three scenarios. Natural history shows a gradual increase in risk over time. “EET only” represents initial endoscopic eradication therapy (EET) at Year 0 followed by annual endoscopy; if EAC is detected during surveillance, treatment is provided at that time. “EET + maintenance therapy” represents initial EET followed by periodic therapeutic interventions over approximately 5 years, even in the absence of visible lesions, resulting in sustained suppression of risk.

In contrast, individuals with NDBE derive minimal benefit from uniform surveillance, yet a subset remains at elevated risk. For this group, future work should focus on developing risk-stratified surveillance approaches using molecular biomarkers, AI-enhanced detection, and non-endoscopic technologies. Incorporating individualized risk factors such as smoking, obesity, chronic gastroesophageal reflux, and genetic susceptibility may further improve precision in identifying high-risk NDBE phenotypes who warrant intensified monitoring or early intervention.

### 4.7 Conclusions

Dysplasia-stratified management is essential for optimizing surveillance and treatment strategies in BE. Most individuals with NDBE do not require routine surveillance, whereas any degree of dysplasia warrants EET followed by targeted post-treatment monitoring. These findings underscore the central role of phenotype-specific EAC risk in guiding management decisions and highlight the inefficiency of applying uniform surveillance intervals across heterogeneous risk groups. This paradigm shift has important implications for U.S. practice, moving from long-term surveillance in low-risk individuals toward consistent implementation of EET for LGD and HGD.

## List of Abbreviations

AI: Artificial intelligence
BE: Barrett’s esophagus
CI: Confidence interval
CPT: Current Procedural Terminology
EAC: Esophageal adenocarcinoma
EET: Endoscopic eradication therapy
HGD: High-grade dysplasia
ICER: Incremental cost-effectiveness ratio
LGD: Low-grade dysplasia
miRNA: MicroRNA
NDBE: Nondysplastic Barrett’s esophagus
NMB: Net monetary benefit
OR: Odds ratio
PSA: Probabilistic sensitivity analysis
QALY: Quality-adjusted life-year
SEER: Surveillance, Epidemiology, and End Results
SSBE: Short-segment Barrett’s esophagus
WTP: Willingness-to-pay

## Data Availability

All data used in this study are publicly available aggregated national statistics obtained from publicly accessible U.S. databases, including the Surveillance, Epidemiology, and End Results (SEER) Program. No individual-level data were used.

https://www.cms.gov/medicare/physician-fee-schedule/search

https://seer.cancer.gov/

https://www.cms.gov/medicare/medicare-fee-for-service-part-b-drugs/mcrpartbdrugavgsalesprice

## Acknowledgments

None.

---

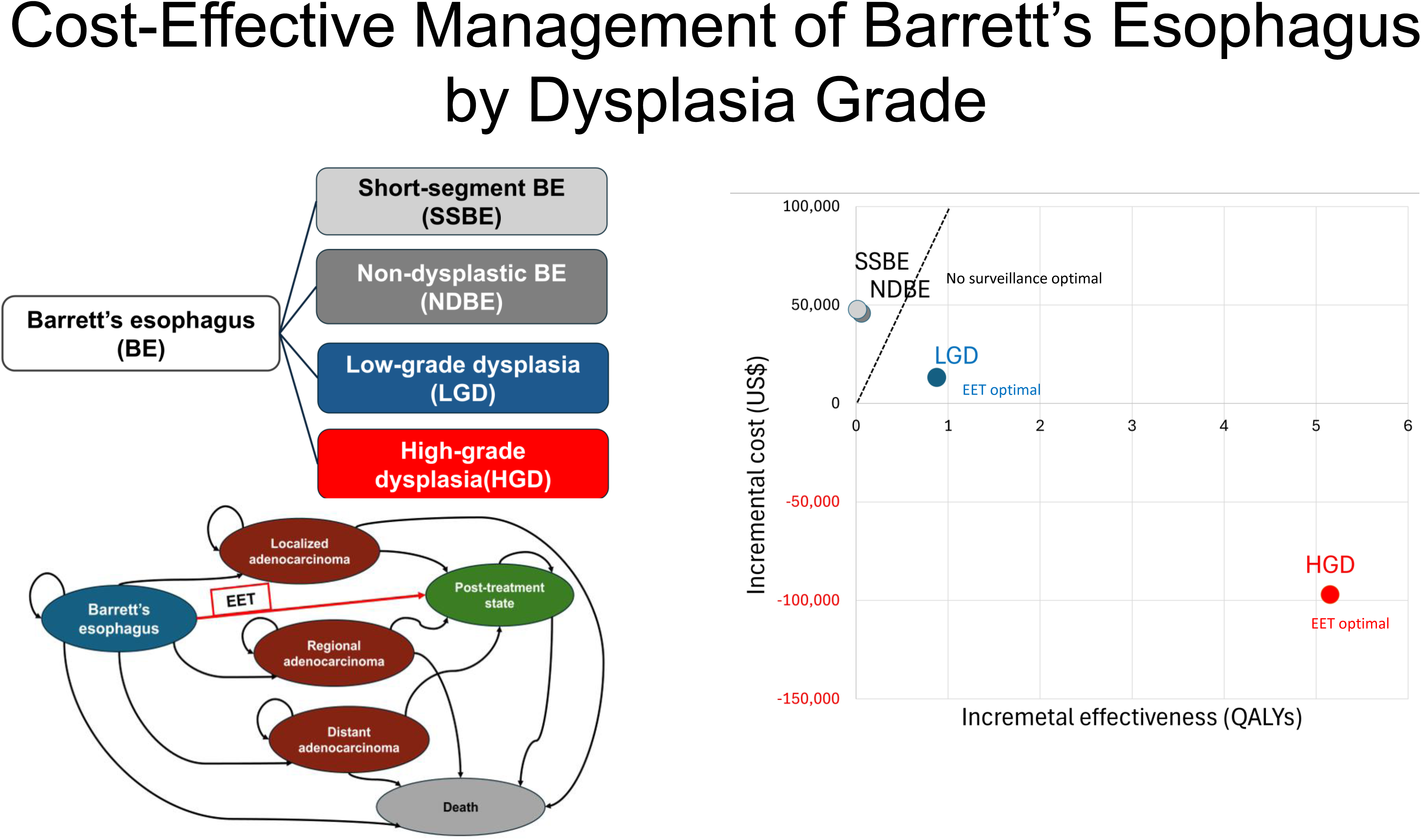

## Notes

### Competing Interest Statement

The authors have declared no competing interest.

### Author Declarations

The study used ONLY openly available aggregated human data that were originally located at publicly accessible U.S. national databases, including the Surveillance, Epidemiology, and End Results (SEER) Program and other publicly available national statistics sources. No individual-level data were used.

