## Supplementary Material for "Dysplasia-Stratified Management of Barrett’s Esophagus: An Incidence-Based U.S. Cost-Effectiveness Analysis"

### Supplementary Table of Contents

**Supplementary Table S1.** Model Inputs and Parameter Values

**Supplementary Table S2.** Base-case results in the SSBE cohort

**Supplementary Table S3.** Base-case results in the NDBE cohort

**Supplementary Table S4.** Base-case results in the LGD cohort

**Supplementary Table S5.** Base-case results in the HGD cohort

**Table S1.** Model Inputs and Parameter Values

| Variable | Baseline value | One-way sensitivity analysis range | | Distribution | Reference |
|---|---|---|---|---|---|
| | | Min | Max | | |
| **Probabilities** | | | | | |
| Annual incidence of EAC in SSBE | 0.0006 | 0.0001 | 0.0010 | Beta | 17 |
| Annual incidence of EAC in NDBE | 0.0021 | 0.0012 | 0.0029 | Beta | 6 |
| Annual incidence of EAC in LGD | 0.0116 | 0.0057 | 0.0175 | Beta | 6 |
| Annual incidence of EAC in HGD | 0.1416 | 0.1243 | 0.1589 | Beta | 6 |
| Dysplasia recurrence after EET in LGD | 0.0600 | 0.0400 | 0.1000 | Beta | 18 |
| EAC recurrence after EET in LGD | 0.0050 | 0.0020 | 0.0100 | Beta | 19 |
| Dysplasia recurrence after EET in HGD | 0.0500 | 0.0300 | 0.1000 | Beta | 18 |
| EAC recurrence after EET in HGD | 0.0100 | 0.0050 | 0.0200 | Beta | 19 |
| Adherence rates | | | | | |
| Endoscopy | 0.78 | 0.50 | 1.00 | Beta | 20 |
| AI-assisted endoscopy | 0.78 | 0.50 | 1.00 | Beta | 20 |
| Sponge test | 0.75 | 0.50 | 1.00 | Beta | 20 |
| Breath test | 0.95 | 0.50 | 1.00 | Beta | Assumption |
| miRNA test | 0.95 | 0.50 | 1.00 | Beta | Assumption |

| | | | | | |
|---|---|---|---|---|---|
| EET | 0.90 | 0.50 | 1.00 | Beta | Assumption |
| **Sensitivity** | | | | | |
| Endoscopy | 0.75 | 0.70 | 0.80 | Beta | 9 |
| AI-assisted endoscopy | 0.93 | 0.88 | 1.00 | Beta | 12 |
| Sponge test | 0.81 | 0.76 | 0.85 | Beta | 13 |
| Breath test | 0.91 | 0.84 | 0.95 | Beta | 14 |
| miRNA test | 0.96 | 0.90 | 0.98 | Beta | 15 |
| **Specificity** | | | | | |
| Endoscopy | 0.70 | 0.60 | 0.80 | Beta | 9 |
| AI-assisted endoscopy | 0.78 | 0.53 | 0.94 | Beta | 12 |
| Sponge test | 0.90 | 0.82 | 0.93 | Beta | 13 |
| Breath test | 0.74 | 0.69 | 0.79 | Beta | 14 |
| miRNA test | 0.95 | 0.87 | 0.98 | Beta | 15 |
| Stage distribution for non-screen-detected EACs | Localized: 0.24, Regional: 0.34, Distant: 0.42 | – | – | Dirichlet | SEER Stat |
| Stage distribution for screen-detected EACs | Localized: 0.60, Regional: 0.30, Distant: 0.10 | – | – | Dirichlet | SEER Stat |
| Stage-specific 5-year survival rates for EAC | Localized: 0.468, Regional: 0.233, Distant: 0.043 | – | – | Beta | SEER Stat |
| Discount rate | 0.03 | 0.02 | 0.05 | Fixed | 16 |

| | | | | | |
|---|---|---|---|---|---|
| **Costs (US$)** | | | | | |
| Endoscopy | 2,500 | 1,250 | 5,000 | Gamma | 21 |
| AI-assisted endoscopy | 3,000 | 1,500 | 6,000 | Gamma | Assumption |
| Sponge test | 400 | 200 | 1,600 | Gamma | 22 |
| Breath test | 200 | 100 | 800 | Gamma | 14 |
| miRNA test | 300 | 150 | 1,200 | Gamma | Assumption |
| Localized EAC treatment | 50,000 | 25,000 | 200,000 | Gamma | 23 |
| Regional EAC treatment | 100,000 | 50,000 | 400,000 | Gamma | 23 |
| Distant EAC treatment | 150,000 | 75,000 | 600,000 | Gamma | 23 |
| EET | 7,000 | 3,500 | 14,000 | Gamma | 24 |
| **Health state utilities** | | | | | |
| SSBE | 0.93 | 0.88 | 0.96 | Beta | 25 |
| NDBE | 0.93 | 0.88 | 0.96 | Beta | 25 |
| LGD | 0.88 | 0.82 | 0.92 | Beta | 25 |
| HGD | 0.82 | 0.75 | 0.88 | Beta | 25 |
| Localized EAC | 0.75 | 0.70 | 0.80 | Beta | 25 |
| Regional EAC | 0.65 | 0.60 | 0.70 | Beta | 25 |
| Distant EAC | 0.55 | 0.45 | 0.60 | Beta | 25 |
| Post-treatment state in EET year: LGD | 0.83 | 0.77 | 0.87 | Beta | 25 |
| Post-treatment state in EET year: HGD | 0.77 | 0.70 | 0.83 | Beta | 25 |
| Post-treatment state: NDBE | 0.93 | 0.88 | 0.96 | Beta | 25 |
| Post-treatment state: LGD | 0.88 | 0.82 | 0.92 | Beta | 25 |

| | | | | | |
|---|---|---|---|---|---|
| Post-treatment state: HGD | 0.82 | 0.75 | 0.88 | Beta | 25 |
| Death | 0 | N/A | N/A | - | - |

AI, artificial intelligence; EAC, esophageal adenocarcinoma; EET, endoscopic eradication therapy; NDBE, non-dysplastic Barrett's esophagus; LGD, low-grade dysplasia; HGD, high-grade dysplasia; SSBE, short-segment Barrett's esophagus; miRNA, microRNA; N/A, not applicable.

**Table S2.** Base-case results in the SSBE cohort

| Strategy | Cost (US$) | Incremental Cost (US$) | Effectiveness (QALY) | Incremental Effectiveness (QALY) | ICER (US$/QALY) | NMB (US$) |
|---|---|---|---|---|---|---|
| No surveillance | 3,053 | Ref | 18.1703 | Ref | Ref | 1,813,980 |
| Sponge test every 10 years | 9,055 | 6,002 | 18.1717 | 0.0014 | 4,290,967 | 1,808,118 |
| Endoscopy every 10 years | 8,824 | -231 | 18.1721 | 0.0004 | dominant | 1,808,389 |
| Breath test every 10 years | 11,854 | 3,030 | 18.1723 | 0.0002 | 15,570,821 | 1,805,379 |
| miRNA test every 10 years | 11,003 | -851 | 18.1724 | 0.0001 | dominant | 1,806,240 |
| AI-assisted endoscopy every 10 years | 9,977 | -1,026 | 18.1726 | 0.0001 | dominant | 1,807,279 |
| Sponge test every 5 years | 12,952 | 2,975 | 18.1727 | 0.0002 | 17,083,609 | 1,804,322 |
| Sponge test every 4 years | 14,907 | 1,956 | 18.1732 | 0.0005 | 3,866,729 | 1,802,417 |
| Endoscopy every 5 years | 12,569 | -2,338 | 18.1734 | 0.0002 | dominant | 1,804,772 |
| Breath test every 5 years | 17,567 | 4,998 | 18.1738 | 0.0003 | 14,962,947 | 1,799,808 |
| miRNA test every 5 years | 16,163 | -1,404 | 18.1739 | 0.0002 | dominant | 1,801,231 |
| Endoscopy every 4 years | 14,449 | -1,714 | 18.1741 | 0.0001 | dominant | 1,802,957 |
| Sponge test every 3 years | 18,171 | 3,722 | 18.1741 | 0.0000 | 191,584,215 | 1,799,238 |
| AI-assisted endoscopy every 5 years | 14,470 | -3,701 | 18.1742 | 0.0001 | dominant | 1,802,945 |
| Breath test every 4 years | 20,434 | 5,964 | 18.1745 | 0.0003 | 19,007,806 | 1,797,013 |

| | | | | | | |
|---|---|---|---|---|---|---|
| miRNA test every 4 years | 18,753 | -1,681 | 18.1747 | 0.0002 | dominant | 1,798,717 |
| AI-assisted endoscopy every 4 years | 16,725 | -2,028 | 18.1750 | 0.0003 | dominant | 1,800,771 |
| Endoscopy every 3 years | 17,586 | 861 | 18.1752 | 0.0002 | 4,555,323 | 1,799,929 |
| Breath test every 3 years | 25,220 | 7,634 | 18.1757 | 0.0005 | 14,627,844 | 1,792,347 |
| Sponge test every 2 years | 24,709 | -511 | 18.1758 | 0.0001 | dominant | 1,792,869 |
| miRNA test every 3 years | 23,075 | -1,634 | 18.1760 | 0.0002 | dominant | 1,794,521 |
| AI-assisted endoscopy every 3 years | 20,489 | -2,586 | 18.1763 | 0.0003 | dominant | 1,797,142 |
| Endoscopy every 2 years | 23,871 | 3,382 | 18.1773 | 0.0010 | 3,326,322 | 1,793,862 |
| Breath test every 2 years | 34,807 | 10,936 | 18.1781 | 0.0008 | 14,441,621 | 1,783,002 |
| miRNA test every 2 years | 31,734 | -3,073 | 18.1785 | 0.0004 | dominant | 1,786,117 |
| AI-assisted endoscopy every 2 years | 28,029 | -3,705 | 18.1790 | 0.0005 | dominant | 1,789,872 |
| Annual sponge test | 44,327 | 16,298 | 18.1809 | 0.0019 | 8,762,766 | 1,773,760 |
| Annual endoscopy | 42,727 | -1,599 | 18.1839 | 0.0030 | dominant | 1,775,658 |
| Annual breath test | 63,572 | 20,844 | 18.1853 | 0.0015 | 14,234,732 | 1,754,960 |
| Annual miRNA test | 57,715 | -5,857 | 18.1861 | 0.0008 | dominant | 1,760,899 |
| Annual AI-assisted endoscopy | 50,651 | -7,063 | 18.1871 | 0.0010 | dominant | 1,768,058 |

“Dominant” was defined as an intervention that is both more effective and less costly than the comparator.

**Table S3.** Base-case results in the NDBE cohort

| Strategy | Cost (US$) | Incremental Cost (US$) | Effectiveness (QALY) | Incremental Effectiveness (QALY) | ICER (US$/QALY) | NMB (US$) |
|---|---|---|---|---|---|---|
| No surveillance | 10,472 | Ref | 17.9005 | Ref | Ref | 1,779,581 |
| Sponge test every 10 years | 16,328 | 5,856 | 17.9054 | 0.0048 | 1,210,048 | 1,774,209 |
| Endoscopy every 10 years | 16,081 | -247 | 17.9067 | 0.0014 | dominant | 1,774,594 |
| Breath test every 10 years | 19,061 | 2,980 | 17.9074 | 0.0007 | 4,427,352 | 1,771,681 |
| miRNA test every 10 years | 18,216 | -845 | 17.9078 | 0.0004 | dominant | 1,772,564 |
| AI-assisted endoscopy every 10 years | 17,198 | -1,019 | 17.9082 | 0.0004 | dominant | 1,773,626 |
| Sponge test every 5 years | 20,089 | 2,891 | 17.9088 | 0.0006 | 4,970,099 | 1,770,793 |
| Sponge test every 4 years | 21,978 | 1,888 | 17.9106 | 0.0017 | 1,085,300 | 1,769,078 |
| Endoscopy every 5 years | 19,680 | -2,298 | 17.9112 | 0.0006 | dominant | 1,771,438 |
| Breath test every 5 years | 24,578 | 4,899 | 17.9123 | 0.0012 | 4,250,268 | 1,766,654 |
| miRNA test every 5 years | 23,188 | -1,390 | 17.9130 | 0.0006 | dominant | 1,768,109 |
| Endoscopy every 4 years | 21,486 | -1,702 | 17.9134 | 0.0004 | dominant | 1,769,855 |
| Sponge test every 3 years | 25,129 | 3,643 | 17.9135 | 0.0001 | 60,789,023 | 1,766,217 |
| AI-assisted endoscopy every 5 years | 21,512 | -3,617 | 17.9137 | 0.0003 | dominant | 1,769,860 |
| Breath test every 4 years | 27,348 | 5,836 | 17.9148 | 0.0011 | 5,427,821 | 1,764,132 |

| | | | | | | |
|---|---|---|---|---|---|---|
| miRNA test every 4 years | 25,685 | -1,664 | 17.9156 | 0.0008 | dominant | 1,765,874 |
| AI-assisted endoscopy every 4 years | 23,679 | -2,006 | 17.9165 | 0.0009 | dominant | 1,767,971 |
| Endoscopy every 3 years | 24,502 | 823 | 17.9171 | 0.0006 | 1,280,576 | 1,767,212 |
| Breath test every 3 years | 31,972 | 7,470 | 17.9189 | 0.0018 | 4,152,384 | 1,759,922 |
| Sponge test every 2 years | 31,443 | -528 | 17.9193 | 0.0004 | dominant | 1,760,486 |
| miRNA test every 3 years | 29,851 | -1,592 | 17.9200 | 0.0007 | dominant | 1,762,144 |
| AI-assisted endoscopy every 3 years | 27,294 | -2,557 | 17.9211 | 0.0012 | dominant | 1,764,818 |
| Endoscopy every 2 years | 30,543 | 3,249 | 17.9246 | 0.0035 | 930,494 | 1,761,919 |
| Breath test every 2 years | 41,234 | 10,691 | 17.9272 | 0.0026 | 4,097,781 | 1,751,489 |
| miRNA test every 2 years | 38,198 | -3,036 | 17.9287 | 0.0015 | dominant | 1,754,672 |
| AI-assisted endoscopy every 2 years | 34,537 | -3,661 | 17.9304 | 0.0017 | dominant | 1,758,503 |
| Annual sponge test | 50,390 | 15,853 | 17.9368 | 0.0064 | dominant | 1,743,288 |
| Annual endoscopy | 48,670 | -1,720 | 17.9471 | 0.0103 | dominant | 1,746,038 |
| Annual breath test | 69,026 | 20,356 | 17.9521 | 0.0050 | 4,037,150 | 1,726,186 |
| Annual miRNA test | 63,243 | -5,783 | 17.9550 | 0.0028 | dominant | 1,732,252 |
| Annual AI-assisted endoscopy | 56,271 | -6,972 | 17.9582 | 0.0033 | dominant | 1,739,554 |

**Table S4.** Base-case results in the LGD cohort

| Strategy | Cost (US$) | Incremental Cost (US$) | Effectiveness (QALY) | Incremental Effectiveness (QALY) | ICER (US$/QALY) | NMB (US$) |
|---|---|---|---|---|---|---|
| No surveillance | 51,169 | Ref | 15.5039 | Ref | Ref | 1,499,217 |
| Sponge test every 10 years | 56,196 | 5,027 | 15.5278 | 0.0239 | 210,083 | 1,496,583 |
| Endoscopy every 10 years | 55,858 | -338 | 15.5346 | 0.0068 | dominant | 1,497,601 |
| Breath test every 10 years | 58,559 | 2,701 | 15.5379 | 0.0033 | 811,519 | 1,495,233 |
| miRNA test every 10 years | 57,748 | -811 | 15.5398 | 0.0019 | dominant | 1,496,231 |
| AI-assisted endoscopy every 10 years | 56,772 | -976 | 15.5420 | 0.0022 | dominant | 1,497,424 |
| Sponge test every 5 years | 59,203 | 2,431 | 15.5442 | 0.0022 | 1,081,678 | 1,495,218 |
| Sponge test every 4 years | 60,716 | 1,513 | 15.5525 | 0.0083 | 182,104 | 1,494,536 |
| Endoscopy every 5 years | 58,641 | -2,075 | 15.5557 | 0.0031 | dominant | 1,496,926 |
| Breath test every 5 years | 62,983 | 4,342 | 15.5613 | 0.0056 | 773,789 | 1,493,145 |
| miRNA test every 5 years | 61,672 | -1,311 | 15.5644 | 0.0032 | dominant | 1,494,772 |
| Endoscopy every 4 years | 60,041 | -1,631 | 15.5663 | 0.0019 | dominant | 1,496,593 |
| Sponge test every 3 years | 63,243 | 3,202 | 15.5664 | 0.0001 | 41,863,092 | 1,493,398 |
| AI-assisted endoscopy every 5 years | 60,093 | -3,150 | 15.5681 | 0.0017 | dominant | 1,496,717 |
| Breath test every 4 years | 65,209 | 5,116 | 15.5731 | 0.0050 | 1,022,548 | 1,492,101 |

| | | | | | | |
|---|---|---|---|---|---|---|
| miRNA test every 4 years | 63,646 | -1,563 | 15.5769 | 0.0038 | dominant | 1,494,045 |
| AI-assisted endoscopy every 4 years | 61,764 | -1,882 | 15.5813 | 0.0044 | dominant | 1,496,369 |
| Endoscopy every 3 years | 62,379 | 615 | 15.5842 | 0.0028 | 216,345 | 1,496,038 |
| Breath test every 3 years | 68,926 | 6,547 | 15.5929 | 0.0087 | 752,586 | 1,490,361 |
| Sponge test every 2 years | 68,306 | -620 | 15.5943 | 0.0014 | dominant | 1,491,119 |
| miRNA test every 3 years | 66,942 | -1,364 | 15.5978 | 0.0035 | dominant | 1,492,834 |
| AI-assisted endoscopy every 3 years | 64,554 | -2,388 | 15.6034 | 0.0057 | dominant | 1,495,790 |
| Endoscopy every 2 years | 67,064 | 2,510 | 15.6199 | 0.0165 | 152,304 | 1,494,928 |
| Breath test every 2 years | 76,374 | 9,310 | 15.6325 | 0.0126 | 740,525 | 1,486,875 |
| miRNA test every 2 years | 73,548 | -2,826 | 15.6396 | 0.0071 | dominant | 1,490,408 |
| AI-assisted endoscopy every 2 years | 70,145 | -3,402 | 15.6478 | 0.0082 | dominant | 1,494,632 |
| Annual sponge test | 83,504 | 13,358 | 15.6779 | 0.0302 | 442,987 | 1,484,289 |
| Annual endoscopy | 81,127 | -2,377 | 15.7274 | 0.0494 | dominant | 1,491,609 |
| Annual breath test | 98,731 | 17,605 | 15.7516 | 0.0242 | 727,146 | 1,476,425 |
| Annual miRNA test | 93,376 | -5,355 | 15.7652 | 0.0136 | dominant | 1,483,141 |
| Annual AI-assisted endoscopy | 86,929 | -6,447 | 15.7810 | 0.0158 | dominant | 1,491,170 |
| EET followed by annual endoscopy | 64,234 | -22,696 | 16.3790 | 0.5980 | dominant | 1,573,669 |

**Table S5.** Base-case results in the HGD cohort

| Strategy | Cost (US$) | Incremental Cost (US$) | Effectiveness (QALY) | Incremental Effectiveness (QALY) | ICER (US$/QALY) | NMB (US$) |
|---|---|---|---|---|---|---|
| No surveillance | 212,485 | Ref | 7.6641 | Ref | Ref | 553,930 |
| Sponge test every 10 years | 212,785 | 300 | 7.8359 | 0.1717 | 1,748 | 570,804 |
| Endoscopy every 10 years | 211,780 | -1,004 | 7.8847 | 0.0488 | dominant | 576,685 |
| Sponge test every 5 years | 212,510 | 729 | 7.8971 | 0.0125 | 58,513 | 577,203 |
| Breath test every 10 years | 213,058 | 548 | 7.9085 | 0.0114 | 47,984 | 577,797 |
| miRNA test every 10 years | 212,338 | -720 | 7.9220 | 0.0134 | dominant | 579,859 |
| Sponge test every 4 years | 212,360 | 22 | 7.9310 | 0.0090 | 2,416 | 580,739 |
| AI-assisted endoscopy every 10 years | 211,481 | -879 | 7.9376 | 0.0066 | dominant | 582,277 |
| Endoscopy every 5 years | 211,129 | -352 | 7.9633 | 0.0257 | dominant | 585,199 |
| Sponge test every 3 years | 212,104 | 975 | 7.9890 | 0.0257 | 37,886 | 586,798 |
| Breath test every 5 years | 212,706 | 602 | 7.9957 | 0.0067 | 90,362 | 586,862 |
| Endoscopy every 4 years | 210,774 | -1,932 | 8.0068 | 0.0111 | dominant | 589,902 |
| miRNA test every 5 years | 211,762 | 988 | 8.0139 | 0.0071 | 138,274 | 589,628 |
| AI-assisted endoscopy every 5 years | 210,637 | -1,125 | 8.0351 | 0.0212 | dominant | 592,870 |
| Breath test every 4 years | 212,515 | 1,878 | 8.0439 | 0.0088 | 213,296 | 591,872 |

| | | | | | | |
|---|---|---|---|---|---|---|
| miRNA test every 4 years | 211,448 | -1,067 | 8.0647 | 0.0209 | dominant | 595,026 |
| Endoscopy every 3 years | 210,168 | -1,279 | 8.0813 | 0.0165 | dominant | 597,958 |
| AI-assisted endoscopy every 4 years | 210,177 | 9 | 8.0890 | 0.0077 | 1,152 | 598,721 |
| Sponge test every 2 years | 211,583 | 1,406 | 8.1075 | 0.0185 | 75,808 | 599,169 |
| Breath test every 3 years | 212,188 | 605 | 8.1264 | 0.0189 | 31,954 | 600,457 |
| miRNA test every 3 years | 210,912 | -1,276 | 8.1519 | 0.0254 | dominant | 604,273 |
| AI-assisted endoscopy every 3 years | 209,393 | -1,518 | 8.1814 | 0.0295 | dominant | 608,744 |
| Endoscopy every 2 years | 208,936 | -457 | 8.2334 | 0.0521 | dominant | 614,406 |
| Breath test every 2 years | 211,522 | 2,586 | 8.2951 | 0.0617 | 41,931 | 617,987 |
| miRNA test every 2 years | 209,821 | -1,701 | 8.3298 | 0.0347 | dominant | 623,155 |
| AI-assisted endoscopy every 2 years | 207,797 | -2,023 | 8.3700 | 0.0403 | dominant | 629,207 |
| Annual sponge test | 210,001 | 2,204 | 8.4680 | 0.0980 | 22,490 | 636,803 |
| Annual endoscopy | 205,195 | -4,807 | 8.6963 | 0.2283 | dominant | 664,437 |
| Annual breath test | 209,501 | 4,306 | 8.8081 | 0.1118 | 38,509 | 671,313 |
| Annual miRNA test | 206,509 | -2,992 | 8.8710 | 0.0629 | dominant | 680,590 |
| Annual AI-assisted endoscopy | 202,953 | -3,556 | 8.9440 | 0.0730 | dominant | 691,451 |
| EET followed by annual endoscopy | 115,450 | -87,503 | 12.8171 | 3.8731 | dominant | 1,166,264 |
